# Laboratory Readiness and genomic surveillance of Covid-19 in the Capital of Brazil

**DOI:** 10.1101/2024.05.10.24307182

**Authors:** Fabrício Vieira Cavalcante, Christina Pacheco Santos Martin, Gustavo Saraiva Frio, Rodrigo Guerino Stabeli, Leonor Maria Pacheco Santos

**Author notes:** These authors contributed equally to this work.

## Abstract

**Objective:** Analyze the diagnostic readiness to Covid-19 and the genomic surveillance of SARS-CoV-2 in Brasília, the capital of Brazil.

**Method:** Retrospective, cross-sectional study, with data from: cases/deaths - Ministry of Health; RT-PCR analyzes Brasília Central Public Health Laboratory (LACEN); genomics - Global Initiative on Sharing All Influenza Data (GISAID).

**Results:** In March 2021, with the Gamma variant being predominant, RT-PCR dosages by LACEN reached their peak, followed by a reduction, possibly due to the start of vaccination. New peaks were observed in September 2021 and January 2022. The average time for releasing RT-PCR results was reduced from eight days (July 2020), to around eight hours in 2023. The participation of private laboratories was evident in sequencing the SARS-CoV-2 variants of concern in Brasília (n=1,897). LACEN received 571 samples, sequencing 50%. A decrease in the incidence of cases and deaths due to Covid-19 was noted in the years 2022 to 2023, following the national trend.

**Conclusion:** LACEN maintained RT-PCR dosages satisfactorily throughout the period. Regarding the genomic surveillance of SARS-CoV-2, the vast majority of samples were sequenced by private laboratories and the sequencing predicted by LACEN was not covered in its entirety.

## Introduction

Covid-19 emerged in China at the end of 2019 and spread quickly to several countries such as the United States of America and countries in South America [1,2], infecting thousands of people around the world. The World Health Organization (WHO) decided to declare a Public Health Emergency of International Concern (PHEIC) on March 11, 2020, due to the high rate of contagion of SARS-CoV-2 [3]. Currently, Covid-19 is better controlled and the WHO decreed on May 5, 2023 the end of PHEIC regarding SARS-CoV-2 [4].

To achieve control and mitigation of Covid-19, several strategies were necessary, such as the use of a mask, social distancing[5], management of health inputs and financial resources, investment in scientific research[6], treatments, vaccine development[7] and timely diagnosis[8].

Among the measures used to mitigate the disease, is population testing, aiming for early diagnosis and enabling syndromic surveillance, providing support for planning quarantine and other interventions appropriately[9]. The Reverse Transcription Polymerase Chain Reaction (RT-PCR) method is a gold standard test for diagnostic confirmation and for use in public health laboratories[10], being disseminated in several countries as a successful strategy for mitigating Covid-19[11]. In Brazil, the first reports of laboratory diagnosis for SARS-CoV-2 occurred in February 2020, restricted to the National Influenza Center (NIC).The expansion of RT-PCR tests occurred in March 2020, when the NIC trained the all the 27 state Central Health Reference Laboratories (LACENs) and they began to receive the necessary inputs to carry out the diagnostic method[12].

In view of this context and in line with the principles of the Unified Health System (SUS), the Health Care Network (RAS), has the responsibility of the integrality of health actions and services. The LACEN Laboratories assumes its leading role in this epidemiological scenario, since the organizational and operational structure of the RAS is made up of support systems, including diagnostic and therapeutic support[13]. In the meantime, it is understood that diagnostic support permeates all levels of health care and is of important relevance for strengthening the RAS, in order to support the planning of interventions.

Given this, the study aims to analyze the diagnostic response capacity to Covid-19 and the genomic surveillance of SARS-CoV in a timely manner, during the PHEIC period in Brasília, capital city of Brazil.

## Method

This is a retrospective, cross-sectional, exploratory study, based on secondary data in the public domain. It covered the pandemic period, that is, the duration of the Public Health Emergency of International Concern: March 2020 to May 2023.

Public, official data on Covid-19 cases and deaths available on the Ministry of Health portal were considered for the study[14]. Data on vaccination coverage in Brasília were obtained from the Covid-19 Vacinometer portal of the Ministry of Health[15], considering vaccination since the month of January 2021 with a vaccination schedule of second dose or single dose, third dose, booster and additional doses until May 2023. Data from the bivalent vaccine were not included, as there was no specification of which dose was being applied.

Data on diagnostic analyzes were extracted from the InfoSaúde-DF portal of the city of Brasília Health Department[16], including all samples received and analyzed for Covid-19 detection by the RT-PCR method at the Brasília LACEN. The average time for releasing the lab results, in days, was also analyzed.

To identify the dominant variants, throughout the pandemic period, the results of the sequenced genomes of Brasília patients were extracted from the Global Initiative on Sharing All Influenza Data (GISAID) database[17]. The originating laboratories (which provided the samples) and the sending laboratories (which sequenced the genetic material)[18] were also identified. The total number of genomes sequenced was analyzed by originating and sending laboratory, by type of variant and number of cases for Covid-19 for the period from March 2020 to May 2023.

Data consolidation was done in the Excel program (Microsoft Office, 2016). The correlation of the variable ‘time for release of sample results in days’ and the number of samples per month/year, vaccination schedule (primary and first booster) and deaths for the years 2020 to 2023 was analyzed, using the Pearson Correlation Coefficient, in Stata (version 18). Data relating to the SARS-CoV-2 variants of concern (VOC) established by the WHO[19] per sampling period in Brasília were analyzed according to the month of sample collection and their classification: Alpha, Gamma, Delta and Omicron.

As it is secondary data in the public domain, this study does not require approval by the Ethics Committee for research with human beings[20].

## Results

The city of Brasília LACEN Laboratory analyzed a total of 500,789 samples using the RT-PCR method, with 2021 being the year with the highest number of analyzes (n=264,738), followed by 2020 (n=165,500), 2022 (n=58,050) and 2023 (n=12,501). Comparing the number of analyzes using the RT-PCR method with the number of Covid-19 cases in Brasília (Figure 1), in 2020 the number of analyzes for diagnosing Covid-19 increased until July/August and then reduced, returning only when the Alpha variant becomes predominant in Brasília, in December 2020. From January 2021, when the Gamma variant becomes predominant, an oscillation in morbidity due to Covid-19 in Brasília is noted, which is accompanied by a similar oscillation in the number of RT-PCR analyzes in the period. In September 2021, the predominance of the Delta variant was evident, accompanied by an increase in laboratory analyzes and cases for Covid-19, when compared to the previous month (Figure 1).

**Figure 1.**
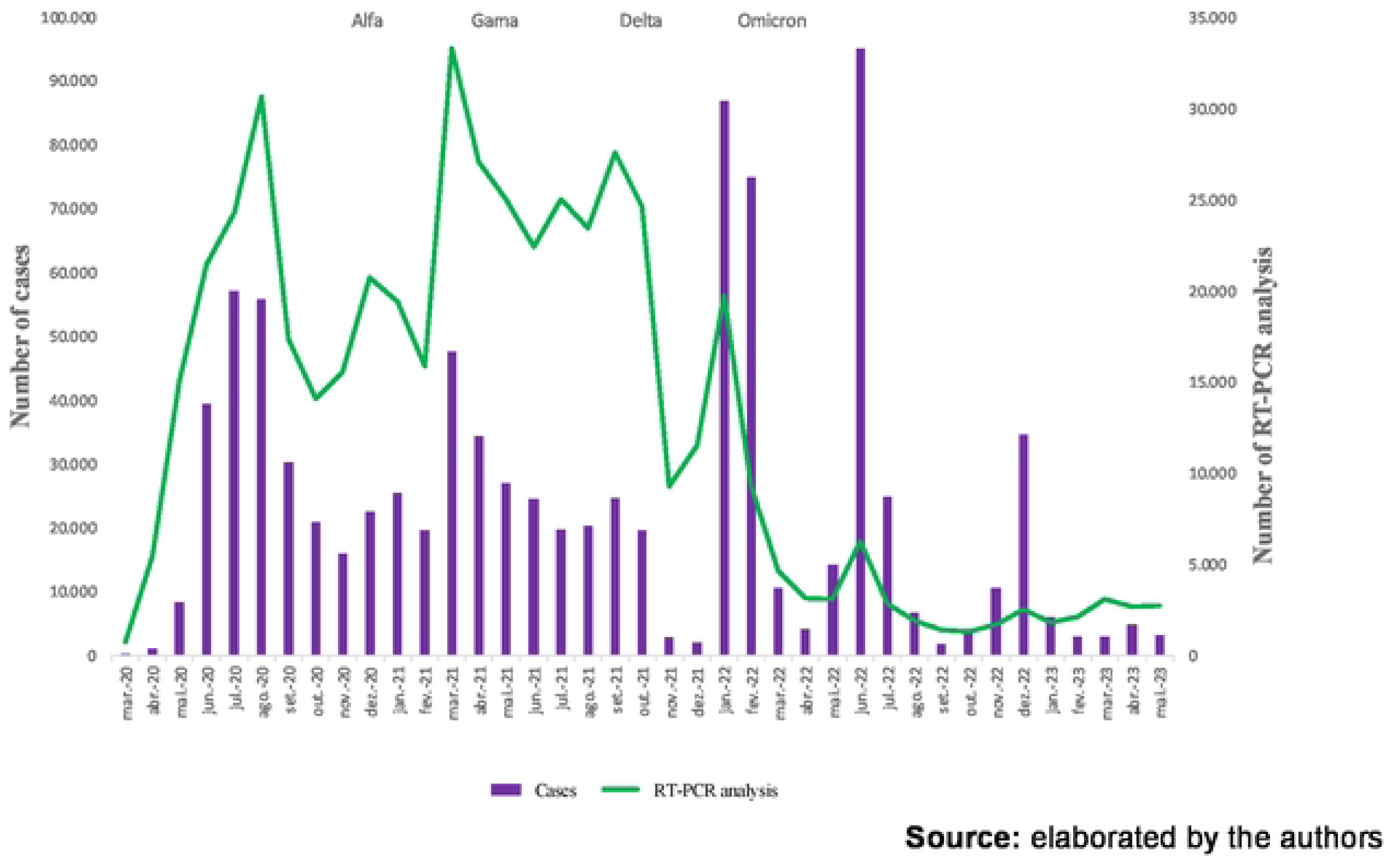
Monthly number of RT-PCR analyzes carried out at the Central Public Health Laboratory and number of Covid-19 cases, compared with the SARS-CoV-2 variant predominant in Brasília, 2020 to 2023.

It is noteworthy that with the predominance of the Omicron variant, the number of cases breaks records in January 2022 (86 thousand cases) and, again, in June of the same year (95 thousand cases) causing a “blackout” in laboratory response capacity in Brasília. This is because the increase in RT-PCR analysis carried out for Covid-19 in these months fell far short of the demand for registered cases, especially in June 2022 (95 thousand cases versus 6 thousand analyzes carried out). Throughout 2023, there was a decrease in analyzes and the number of cases in Brasília (Figure 1), following the national trend.

When observing the number of deaths from Covid-19 in Brasília, it is possible to notice a fluctuation in their number over the three years, with the highest occurrence in April 2021, a period in which the Gamma variant was predominant. The peak of deaths in Brasília occurred in April 2021 (1,769 deaths), precisely one month after the peak of samples analyzed by RT-PCR (33,286 samples). After this date, there was a decrease in the number of deaths from the disease over the months and years, reducing to less than 10 deaths per month from August 2022 onwards. It is worth noting that the laboratory “blackout” occurred in June 2022 was not reflected in an increase in deaths, probably due to the characteristics of the Omicron variant: high transmissibility accompanied by lower lethality, compared to other strains of interest (Figure 2).

**Figure 2.**
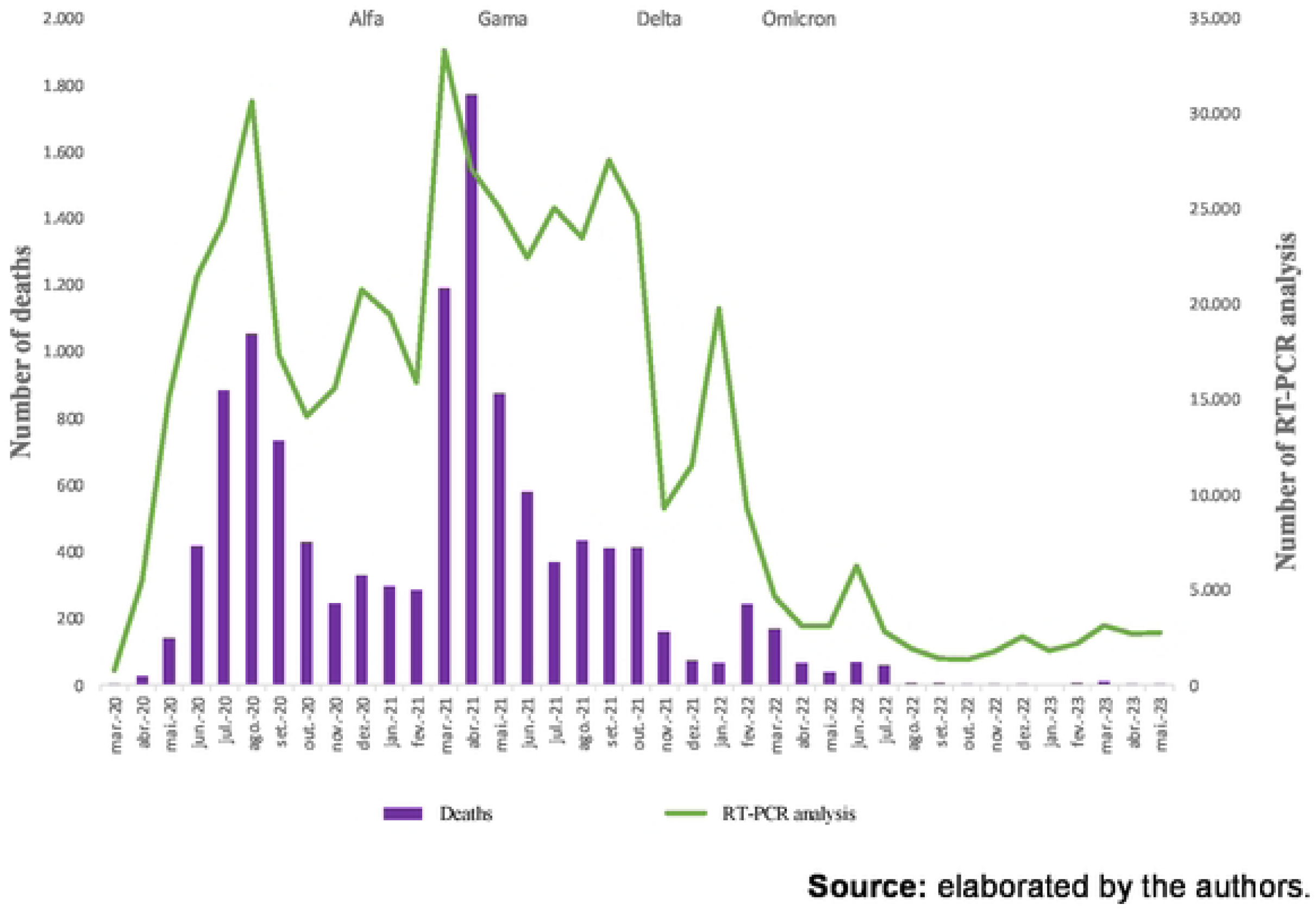
Monthly quantity of RT-PCR analysis carried out at the Central Public Health Laboratory and number of deaths from Covid-19, compared with the SARS-CoV-2 variant predominant in Brasília, 2020 to 2023.

Figure 3 shows the comparison between the number of deaths, of people vaccinated – initial and booster schedule – and the emergence of variants of concern. According to data from the Ministry of Health, the population of Brasília in 2023 was 3,015,268 people. When Brasília reaches 50% of the vaccinated population (September 2021), the number of analyzes reduced significantly, rising again only in the peaks caused by the Omicron variant. The month of September 2021 is when the booster dose begins to be applied, which also contributes to the reduction in the number of deaths in the subsequent months. The peak of deaths, as previously identified, occurred in April 2021, in line with the progress of the vaccination schedule.

**Figure 3.**
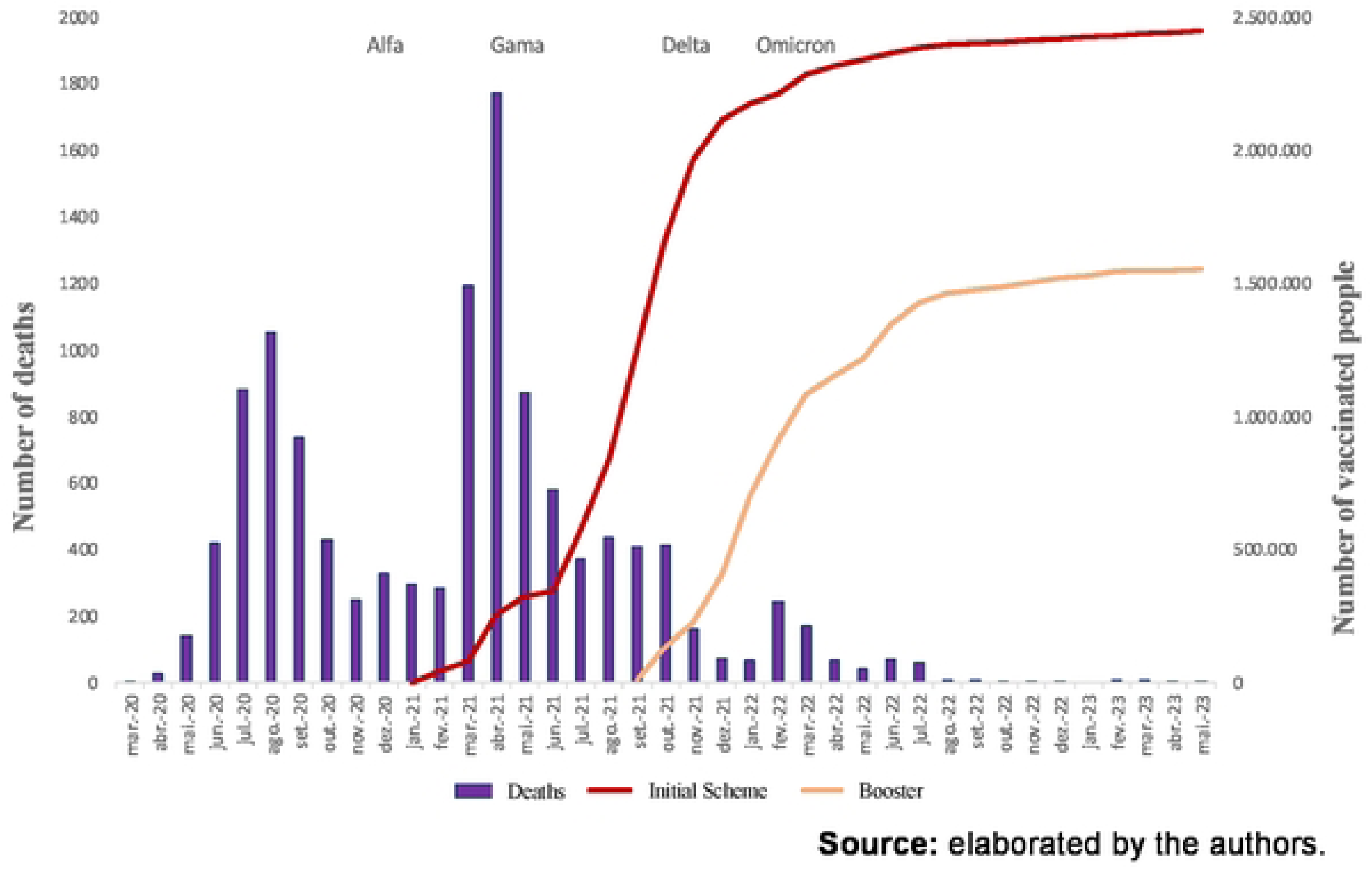
Number of registered deaths, vaccination schedule and predominant SARS-CoV-2 variants in Brasília, 2020 to 2023.

As far as the average time for releasing results by the LACEN, it was longer in the months of July 2020 (around eight days for release), followed by the months of June (three days), May and August, both around two days for the analysis results to be released. It is noteworthy that for the years 2021, 2022 and 2023 the average number of days to release exam results was less than one day, followed by a decrease in January 2021 from 0.99 to 0.28 to obtain the exam result for Covid-19 (Table 1). The average time for releasing RT-PCR results has reduced from up to 8 days in 2020, to around eight hours in 2023.

**Table 1.** Month and average release period in days of the results of RT-PCR analysis from the Central Public Health Laboratory, Brasília, 2020 to 2023.

| Month/Year | 2020 | 2021 | 2022 | 2023 |
| --- | --- | --- | --- | --- |
| Monthly average, in days, for releasing RT-PCR analysis results |  |  |  |  |
| January | . | 0.99 | 0.77 | 0.28 |
| February | . | 0.52 | 0.52 | 0.33 |
| March | 0.79 | 0.94 | 0.39 | 0.35 |
| April | 0.79 | 0.69 | 0.41 | 0.29 |
| May | 2.12 | 0.79 | 0.39 | 0.38 |
| June | 3.89 | 0.95 | 0.47 | . |
| July | 8.33 | 0.57 | 0.45 | . |
| August | 2.51 | 0.45 | 0.35 | . |
| September | 1.03 | 0.92 | 0.30 | . |
| October | 0.69 | 0.77 | 0.28 | . |
| November | 0.62 | 0.44 | 0.23 | . |
| December | 1.34 | 0.43 | 0.34 | . |

**Source:** elaborated by the authors from InfoSaúde-DF<sup>16</sup>

It is important to highlight the correlation between the variables of interest in this work (number of RT-PCR analyses, confirmed cases, deaths, total number of people with the initial vaccination schedule and with the booster dose). There is no significant correlation between the number of cases and vaccination in Brasília.

It should be noted, however, that the number of RT-PCR analyzes for Covid-19 is positively correlated with the number of cases (47.35%) and the number of deaths in the month (80.51%). On the other hand, the advancement of vaccination is negatively correlated with the number of cases: initial scheme (−85.40%) and booster dose (−88.47%). Initial and booster vaccinations are negatively correlated with the number of deaths (−75.99% and −83.71%, respectively). As expected, the Initial Schedule and Booster Dose are highly correlated (90.79%).

Evaluating the provenance of the SARS-CoV-2 sequences available in the GISAID database in Brasília (Figure 4), it is observed that a large part of the samples were sequenced by private laboratories, the vast majority by laboratories from the DASA group (1,897) and a small portion of the samples collected by the Hermes Pardini Institute (43).

**Figure 4.**
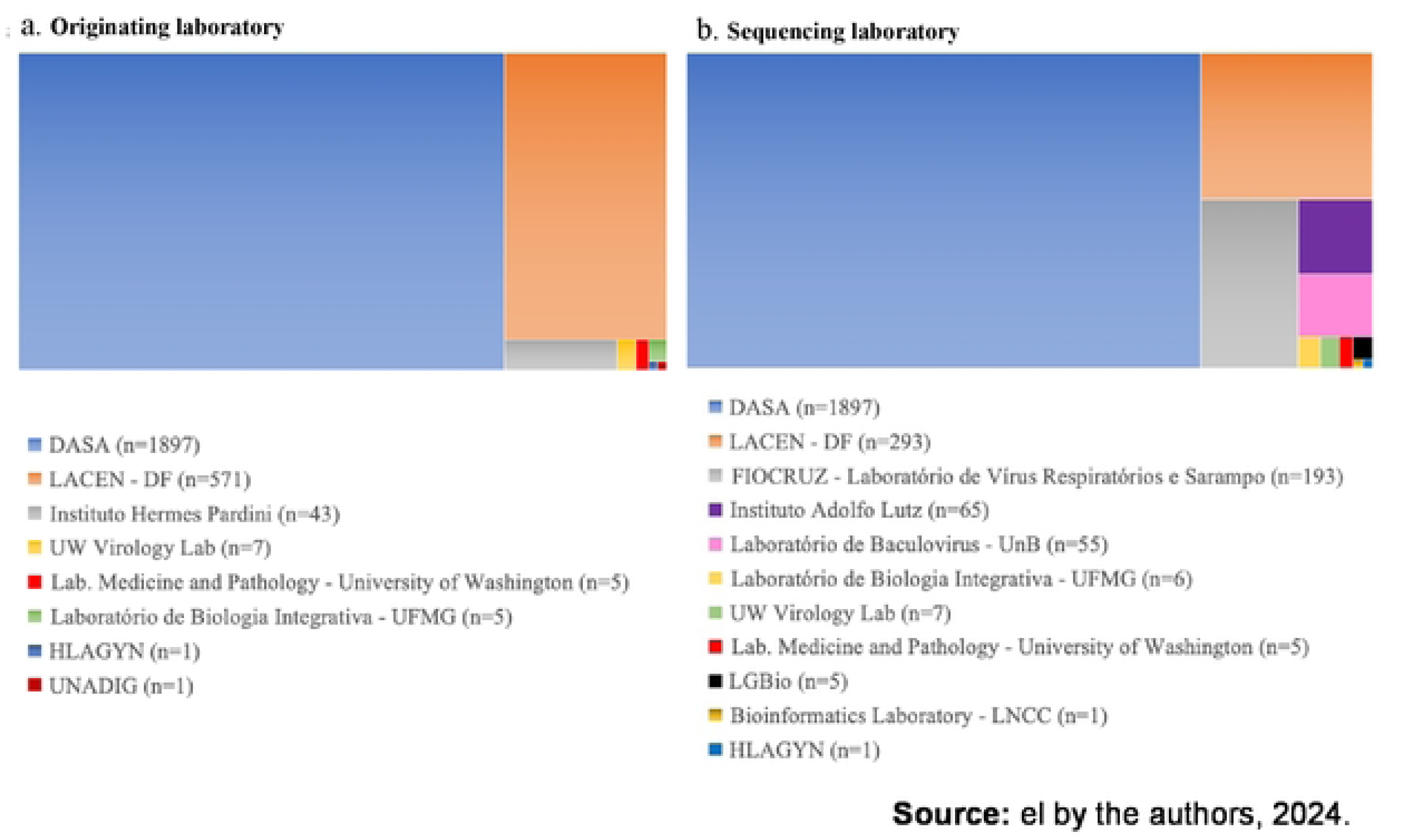
Characterization of the number of Covid-19 samples received and sequenced in public and private national and international laboratories for demand in Brasília from 2020 to 2023.

Among public laboratories, LACEN stands out, which received 571 samples and managed to sequence more than 50% (293). In sequencing the samples, there was significant participation by FIOCRUZ in the Respiratory Virus and Measles Laboratory (193), the Adolfo Lutz Institute (65) and research laboratories at Brazilian public universities such as the University of Brasília – UNB (55), Federal University of Minas Gerais - UFMG (6) and Federal University of Goiás - UFG (5). Some American laboratories, from the University of Washington, also worked on sequencing SARS-CoV-2 samples from the Federal District: Virology Lab (7) and Laboratory Medicine and Pathology (6) (Figure 4).

## Discussion

The data from the present study show an increase in the number of RT-PCR analyzes in Brasília in April 2020, in line with the arrival of the first batch of 10 million tests purchased by the Ministry of Health (MS) to expand coronavirus testing in the country[21], in addition to peaks in analyzes by the Brasília LACEN that coincide with the predominance of the different variants of SARS-CoV-2, with the last identified, and less lethal, Omicron variant. The Omicron variant appears in a scenario of implementation of the vaccination schedule and release of booster doses and in the midst of the third wave of morbidity caused by Covid-19[22].

A decrease in RT-PCR analyzes is evident from September 2021 onwards, in Brasília, possibly due to the introduction of rapid tests in Brazil: some of them only allowed the identification of antibodies (negative or positive result, without specifying the type), while other tests track IgM and IgG antibodies distinctly[23]. These findings are in line with the MS initiative to start rapid testing by antigen test in Brasília through the National Testing Plan for Covid-19 in August 2021[24], and which the following year had its second edition called the National Plan for Expansion of Testing for Covid-19 (PNE – Test), that aimed to expand diagnosis for Covid-19 through rapid antigen testing in symptomatic and asymptomatic individuals based on the care diagnosis strategy, active search and screening[12].

The decrease in RT-PCR analyzes at the Brasília LACEN also comes from Anvisa’s authorization through Collegiate Board Resolution (RDC) n° 377 of April 28, 2020[25], which allows pharmacies and drugstores to carry out rapid tests to detect Covid-19, on an exceptional and temporary basis. These hypotheses are in line with the literature review[26], which describes that serological tests are widely used to mitigate SARS-CoV-2 but points the limitations and advantages in their applicability, which are: the possibility of the method not detecting the disease in asymptomatic patients, in addition to not being adopted exclusively as a parameter for ruling out infection. Among its advantages is the low cost of acquiring the tests when compared to the RT-PCR method, which is evident from the economic analysis study[27] that highlight the advantages of serological tests in relation to the reduction in total costs for health services, ease of use, decentralization in places without laboratory support and test response time. However, a disadvantage is that not all results from these tests were incorporated into the information system centralized at the Ministry of Health, which makes it difficult for the health authority to monitor the evolution of the pandemic from then on. The fact is in line with the limitation evidenced by Leite et al., (2021)[28]: the difficulty in implementing information regarding the performance of laboratories trained to face SARS-CoV-2 in the Regions of the Americas.

Regarding response time, the longest average number of days to release results by Brasília LACEN was eight days in July 2020 when compared to all other months and years analyzed. This lag time is within the time parameters for releasing results within three days, as explained by the Ministry of Health[29] regarding the estimated time for tests to detect Covid-19: rapid antigen test (TR-Ag) with a time of 20 minutes; antigen self-test (AT-Ag) with a time of 20 minutes and the RT-PCR with a time of 72 hours (3 days).

Of the 2,528 Covid-19 samples sequenced from Brasília, only 11.6% were carried out by the Brasilia LACEN, thus requiring the collaboration from other public and private laboratories, teaching and research institutions, accompanied by a global shortage of inputs related to assistance and laboratory diagnosis[12]. Furthermore, the virus mutation process explains the appearance of the Alpha, Gamma, Delta and Omicron variants in Brasília, plus the reinfection of the disease, the decrease in the effectiveness of vaccines and the decrease in sensitivity to diagnostic tests, reinforcing the need to maintain non-pharmacological measures and the acceleration of immunization as a strategy for managing and controlling the circulation of the virus and new mutations[22,30].

This study presents limitations inherent to studies with secondary data, such as the occurrence of inaccuracy and/or incompleteness of data, the use of absolute data on both cases and deaths, which makes it impossible to verify the incidence of the disease and mortality rate, in addition to be an exploratory study without aiming to search for causal results.

It can be seen that Brasilia LACEN response capacity was correlated with the number of cases and deaths due to Covid-19, due to the monitoring of the diagnostic sector in carrying out RT-PCR analyzes that fluctuated during the period analyzed. Despite not having a positive correlation between vaccination and the number of cases and deaths, it is noted that vaccination coverage was associated with a decrease in the transmissibility of the disease and consequently in the number of deaths from Covid-19.

The Brasília LACEN had a decrease in the demand for RT-PCR analyzes in the years 2022 and 2023, however, throughout the respective year it maintained an average of less than one day to release test results in a satisfactory manner based on the time estimated by the Ministry of Health. Regarding the genomic surveillance of SARS-CoV-2 in Brasília, the vast majority of samples were sequenced by private laboratories and those sequenced by Brasilia LACEN were not covered in their entirety for the years 2020 to 2023.

This study showed satisfactory results in the diagnostic readiness to SARS-CoV-2 in Brasília. It discuss and provides strategies that will contribute to government and health entities, both nationally and internationally, to increase laboratory readiness, both to face Covid-19 and other viral health crises that may emerge.

## Data Availability

Where possible, we encourage authors to provide relevant data underlying their research with their submission in order to facilitate editorial and peer review. As far as relevant underlying information about our research, we wish to declare that the submitted manuscript is part of a broad research project, recently approved by the University of Brasília’s Ethical Committee (Annex 1). The main project’s objective is to evaluate the response capacity of the network of 27 Central Public Health Laboratories (LACEN) in Brazil in the context of the Covid-19 pandemic. This effort expect to be able to evaluate the Laboratory Response Capacity in order to prepare them for future epidemics, endemics, outbreaks and pandemics that require an immediate response from the epidemiological and health surveillance sector at a national level. We hope that lessons, applicable to other settings, could be drawn from the Brazilian Unified Health System’s experience (SUS).

